# Tailored Magnetic Resonance Fingerprinting

**DOI:** 10.1101/2022.09.15.22279855

**Authors:** Pavan Poojar, Enlin Qian, Maggie Fung, Patrick Quarterman, Sachin R. Jambawalikar, Angela Lignelli, Sairam Geethanath

**Affiliations:** Accessible Magnetic Resonance Laboratory, Biomedical Imaging and Engineering Institute, Department of Diagnostic, Molecular and Interventional Radiology, Icahn School of Medicine at Mount Sinai, New York, NY, United States; Columbia Magnetic Resonance Research Center, Columbia University in the city of New York, NY, USA; GE Healthcare Applied Sciences Laboratory East, New York, NY, USA; Department of Radiology, Columbia University Irving Medical Center, Columbia University in the city of New York, NY, USA

**Keywords:** Synthetic MRI, MR fingerprinting, Rapid imaging, Multi contrast imaging, Qualitative and quantitative imaging, Brain imaging

## Abstract

Neuroimaging of certain pathologies requires both multi-parametric qualitative and quantitative imaging. The role of the quantitative MRI (qMRI) is well accepted but suffers from long acquisition times leading to patient discomfort, especially in geriatric and pediatric patients. Previous studies show that synthetic MRI can be used in order to reduce the scan time and provide qMRI as well as multi-contrast data. However, this approach suffers from artifacts such as partial volume and flow. In order to increase the scan efficiency (the number of contrasts and quantitative maps acquired per unit time), we designed, simulated, and demonstrated rapid, simultaneous, multi-contrast qualitative (T_1_ weighted, T_1_ fluid attenuated inversion recovery (FLAIR), T_2_ weighted, water, and fat), and quantitative imaging (T_1_ and T_2_ maps) through the approach of tailored MR fingerprinting (TMRF) to cover whole-brain in approximately four minutes.

We performed TMRF on *in vivo* four healthy human brains and *in vitro* ISMRM/NIST phantom and compared with vendor supplied gold standard (GS) and MRF sequences. All scans were performed on a 3T GE Premier system and images were reconstructed offline using MATLAB. The reconstructed qualitative images were then subjected to custom DL denoising and gradient anisotropic diffusion denoising. The quantitative tissue parametric maps were reconstructed using a dense neural network to gain computational speed compared to dictionary matching. The grey matter and white matter tissues in qualitative and quantitative data for the *in vivo* datasets were segmented semi-automatically. The SNR and mean contrasts were plotted and compared across all three methods. The GS images show better SNR in all four subjects compared to MRF and TMRF (GS>TMRF>MRF). The T_1_ and T_2_ values of MRF are relatively overestimated as compared to GS and TMRF. The scan efficiency for TMRF is 1.72 min^-1^ which is higher compared to GS (0.32 min^-1^) and MRF (0.90 min^-1^).

## INTRODUCTION

The value of quantitative MRI (qMRI) in diagnostic medical imaging is well established [1–5]. Clinical studies use qMRI to investigate brain tumors, epilepsy, multiple sclerosis, among other pathologies [6]. However, qMRI requires long acquisition times [7,8]. Moreover, radiologists and technicians need training in specialized software to analyze these qMRI data [9]. Also, MRI protocols always require multi-contrast MR images regardless of qMRI acquisitions [10,11]. Clinical neuroimaging exams such as brain tumor imaging [12], Parkinson’s disease [13], and epilepsy [14,15] utilize both multi-contrast qualitative and quantitative imaging for accurate diagnosis [16]. Independently acquiring weighted images for a multi-contrast exam takes 20 to 30 minutes [17]. These long acquisition times lead to patient discomfort resulting in motion artifacts, especially in geriatric and pediatric populations [18]. Consequently, this long acquisition time reduces the scan efficiency (the number of MR contrasts and quantitative maps acquired per unit time). Hence, there is a need for rapid, simultaneous, multi-contrast, qualitative, and quantitative imaging, which increases the efficiency and, in turn, improves the throughput. Previous solutions to these challenges can be classified into three categories:

### Multi-contrast methods

Examples of accelerated multi-contrast methods include triple contrast rapid acquisition with relaxation enhancement (TCRARE) [19]. This method provides proton density (PD), T_1_, and T_2_ weighted images simultaneously within 2 minutes. Furthermore, parallel imaging [20] and compressed sensing [21–25] reduce acquisition time. Recently, a multi-contrast EPI pulse sequence (EPIMix) for brain MRI provided six contrasts in one minute [26]. However, EPIMix has limitations such as reduced image quality and geometric distortion and thus can be used for screening but cannot replace gold standard sequences [27].

### Rapid quantitative imaging

Methods such as DESPOT1, DESPOT2 [28], quantification of relaxation times and proton density by twin-echo saturation-recovery turbo-field echo (QRAPTEST) [29], quantification of relaxation times and proton density by the multi-echo acquisition of a saturation-recovery using turbo spin-echo readout (QRAPMASTER) [30] accelerate quantitative imaging. However, these methods do not provide weighted images directly from the scanner but can be computed synthetically [31–35]. In contrast, MR fingerprinting (MRF) generates multiple parametric maps simultaneously with high scan efficiency [30,31].

### Synthetic MRI

Synthetic MRI uses parametric maps as inputs to generate multiple weighted images using the MR signal equation [36–38]. A multi-pathway multi-echo acquisition method was developed to acquire 3D multi-parametric maps and generate multi-contrast images using neural networks [39]. In these methods, the images are “synthetically” generated based on MRF and training data rather directly from raw data. Liu et al. [40] have developed a method to generate multi-contrast images and multi-parametric maps simultaneously with the help of a multi-echo gradient echo sequence. However, this method does not provide T_1_, T_2_, or fluid attenuated inversion recovery (FLAIR) contrasts routinely used in clinical settings. Studies show that synthetic MRI has a low image quality for the FLAIR contrast, white noise and flow artifacts [37].

To overcome long acquisition times related to quantitative imaging and the challenges in synthetic MRI, such as the presence of multiple tissue types in one voxel, we introduce tailored magnetic resonance fingerprinting (TMRF). In this work, we design, simulate, and demonstrate simultaneous, natural (non-synthetic), multi-parametric qualitative, and quantitative rapid MR imaging. We accomplish this by tailoring the MRF acquisition schedule (repetition time (TR), echo time (TE), and flip angle (FA)) in approximately four minutes. We also compare TMRF acquired *in vitro* and *in vivo* data with vendor-supplied gold standard (GS) sequences. The multi-contrast images include T_1_ weighted, T_1_ FLAIR, T_2_ weighted, water, and fat. In addition, TMRF provides T_1_ and T_2_ maps.

## MATERIALS AND METHODS

### Framework

The TMRF method involves the design of magnetization evolution, simulation, acquisition, and reconstruction. Figure 1 shows the TMRF framework from simulation to image analysis.

**Figure 1:**
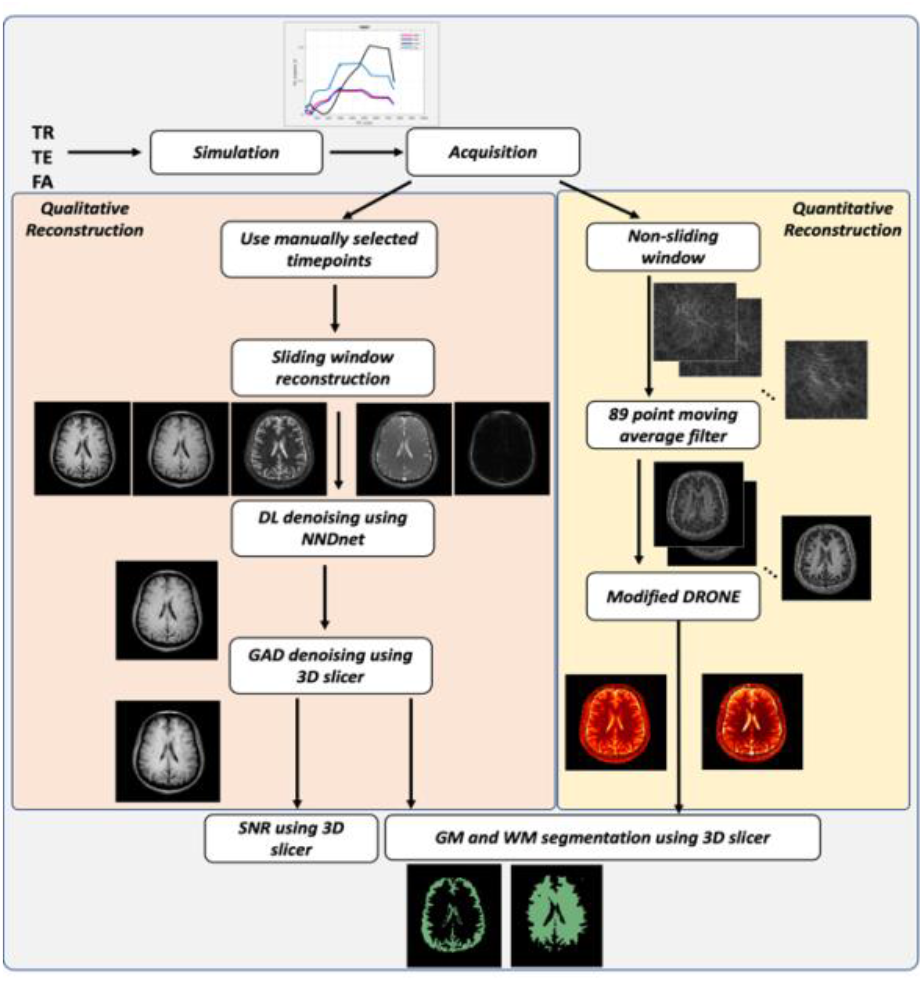
Framework: TMRF framework involves simulation, acquisition, reconstruction, and image analysis. The qualitative (*in vivo*) and quantitative (*in vivo* and *in vitro*) followed two separate pipelines. For qualitative reconstruction, images were reconstructed using manually selected time points. The reconstructed images were then passed through the sliding window with a window size of 89 to get fully sampled k-space. For the first dataset, all 749 time points were used for the sliding window in order to find the time points manually for all the five contrasts and the reconstruction was carried out on only these five time points. This is then followed by image filtering (NNDnet and GAD) and then WM/GM segmentation was performed using 3D Slicer to get SNR and mean intensity values. For the quantitative reconstruction pipeline, first non-sliding window reconstruction was performed followed by 89 points moving average filter. These 749 filtered images were then used to get the T_1_ and T_2_ maps with the help of the modified DRONE method. The WM and GM of *in vivo* quantitative data were segmented to compute T_1_ and T_2_ values with the help of 3D Slicer whereas ROI analysis was used to validate the *in vitro* quantitative data; TMRF - tailored magnetic resonance fingerprinting, NNDnet - native noise denoising network, GAD - gradient anisotropic diffusion, WM - white matter, GM - grey matter, SNR - signal to noise ratio, DRONE - Deep RecOnstruction NEtwork.

### Design and simulation

We designed a steady-state free precession (SSFP) sequence with spiral readouts to contain unique signal evolutions for four different brain tissue matters. FA can modulate MR image contrast derived from such a sequence more than TR [41–43]. MRF has exploited this modulation to significantly vary FA while restricting the TR to a much smaller range above the minimum TR achievable (14.7ms). In this work, magnetization preparation in the form of an inversion pulse was utilized to suppress short and long relaxation components like fat and liquids at different temporal points. This tissue matter-dependent “tailored design” choice allowed for distinct relaxation contrast “windows” and signal constancy in a given contrast window.

TMRF used a total of 749 time points to tailor the magnetization evolution of four tissue types: white matter (WM), grey matter (GM), cerebrospinal fluid (CSF), and fat. We tailored the magnetization evolutions by designing acquisition blocks targeting one or more contrast windows within the block. Each block (250 time points) was designed in this implementation by choosing different FAs (5°, 45°, and 70°) with a minimum TR of 14.7ms. These FA values will be referred to as base FAs for each of the contrast blocks. In each of the three cases, a normally distributed noise with a zero mean and a standard deviation of 0.5 was added to the minimum TR and the base FAs. These two vectors were then sorted in ascending order to avoid spikes in the magnetization evolution. A 90° pulse was introduced after the 250^th^ time point. The TE was held constant at its minimum (1.91ms), except between the 500^th^ and 749^th^ time point (Dixon contrast window [44]). For 2-point Dixon contrast, optimized values of TE_1_ (2.3ms) and TE_2_ (3.4ms) were used to allow the in-phase and out-of-phase acquisition, respectively. Figure 2 shows the TMRF acquisition parameters and the corresponding MRF schedule as in ref. [45]. Extended phase graph [46] was used to simulate the magnetization evolution dictionary for a range of T_1_ (0 to 4000ms in steps of 20ms) and T_2_ (0 to 400ms in steps of 20ms; 450 to 600ms in steps of 50ms; 700ms to 2000ms in steps of 500ms) values.

**Figure 2:**
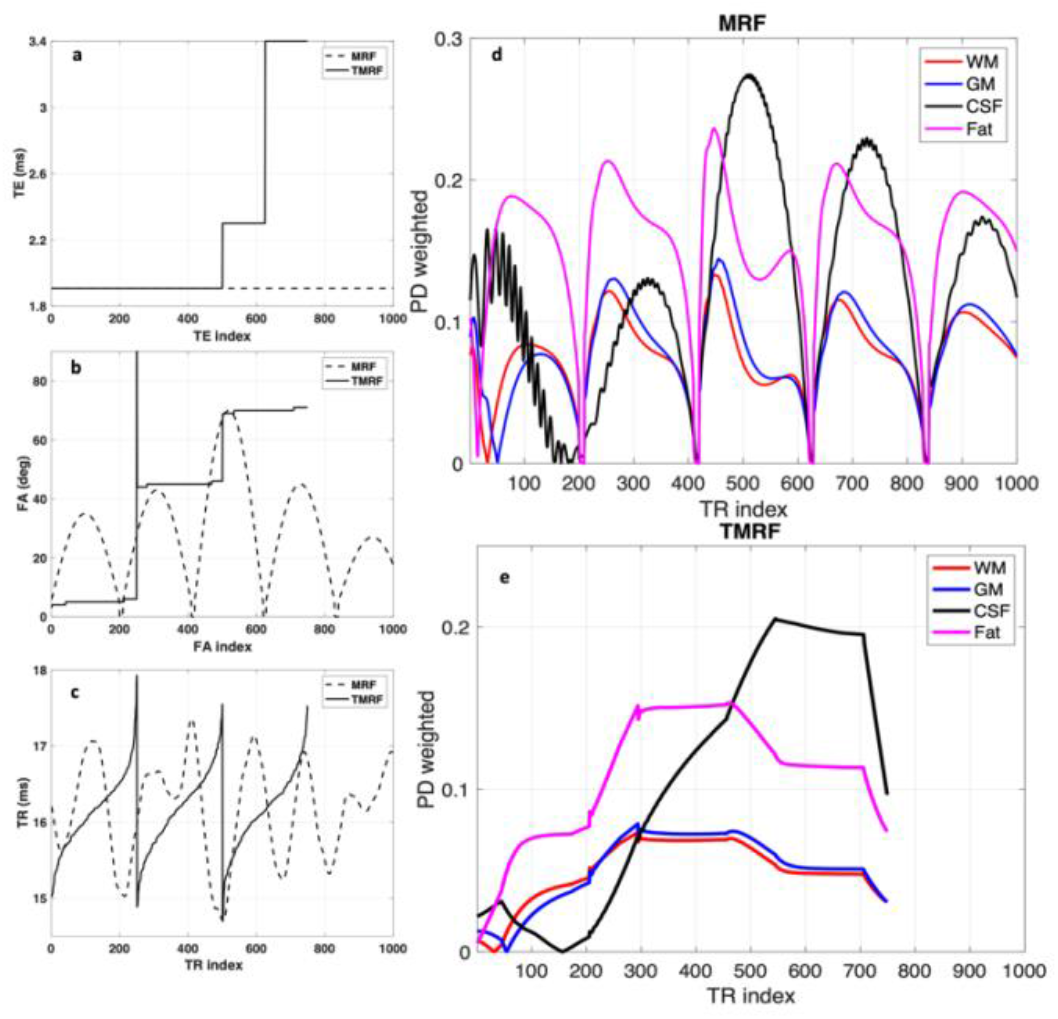
TE, FA, TR and simulation: (a-c) TE, FA, and TR values for the 1000 and 749 time points acquisition for MRF (dashed lines) and TMRF (solid lines) respectively. The TR and FA schemes of TMRF are smoother compared to MRF. This facilitates better sliding window reconstruction. (d) and (e) shows the EPG simulations of the MRF and TMRF acquisition schedules for WM, GM, fat, and CSF respectively. We designed each block to have 250 time points by choosing distinct FA values (5°, 45°, and 70°) including a minimum TR of 14.7ms. A 90° pulse was introduced at the 250^th^ time point to enable subsequent contrasts (water and fat). TE was held at a minimum of 1.91ms, excluding the Dixon contrast window (time points between 500 and 749). For 2-point Dixon contrast, optimized TE values, TE_1_ - 2.3ms and TE_2_ - 3.4ms, were used for in-phase and out-of-phase acquisition, respectively; TE - echo time, FA - flip angle, TR - repetition time, MRF - magnetic resonance fingerprinting, TMRF - tailored MRF, EPG - extended phase graph, WM - white matter, GM - grey matter, CSF - cerebrospinal fluid.

The reconstruction consists of two separate pipelines - qualitative and quantitative. In qualitative reconstruction, all 749 time points (frames) were reconstructed using the sliding window method with a window size of 89 to obtain a fully sampled k-space. The water and fat images were computed using the 2-point Dixon method. All reconstructed images were visually inspected to ensure that all five contrasts were acquired and their corresponding time points were noted. The selected time points were: 1, 95, and 150 for T_2_ weighted, T_1_ FLAIR, and T_1_ weighted, respectively. 2-point Dixon images were derived from time points 575 and 675. This time point selection was performed only for the first dataset. Subsequently, the sliding window reconstruction was performed only on these five time points for the remaining three datasets. This selection reduced the reconstruction time from ∼40 minutes (749 time points) to ∼2 minutes (five time points) for each contrast. The reconstructed images were first denoised using NNDnet [47] and then by gradient anisotropic diffusion (GAD) using a 3D Slicer [48]. SNR and mean intensity of WM and GM were computed on denoised images. For *in vivo* quantitative reconstruction, first, non-sliding window reconstruction was performed (undersampled). Subsequently, 89 points moving average filter was applied to the signal evolutions for all voxels. Finally, we used the modified deep reconstruction network (DRONE) [49] to obtain T_1_ and T_2_ maps. The WM and GM were segmented using 3D Slicer to compute the T_1_ and T_2_ values. The same quantitative reconstruction pipeline was followed for *in vitro* data, except region of interest (ROI) analysis was performed instead of GM and WM segmentation.

### MRI experiments

We imaged the ISMRM/NIST phantom and four healthy human volunteers with the GS, MRF, and TMRF sequences on a 3T GE Premier system (GE Healthcare, USA) using a 21-channel head coil. Table 1 lists the pulse sequences, acquisition parameters, and scan times for GS, MRF, and TMRF. Supplementary Table 1 lists the GS measurements of T_1_ and T_2_ of the ISMRM/NIST phantom. All images were reconstructed offline using MATLAB (The Mathworks. Inc., MA). These acquisitions were part of an IRB-approved study that required written informed consent.

**Table 1:**
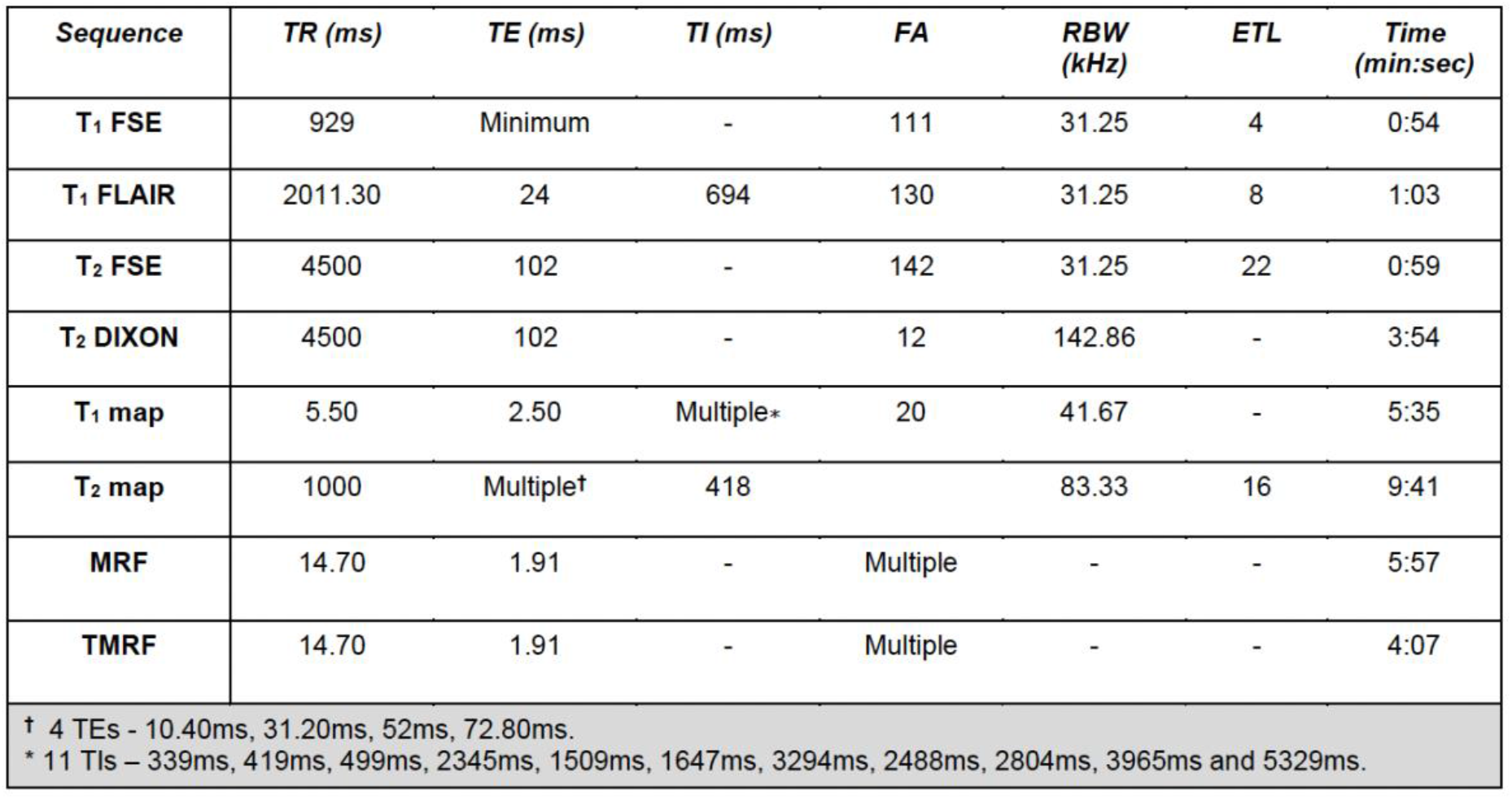
Acquisition parameters: Acquisition parameters for the gold standard, MRF and TMRF sequences. All scans were performed on a 3T GE Premier scanner. For all gold standard sequences, FOV - 22.4×22.4 cm^2^, matrix size-224×224, slices thickness – 5mm, and the number of slices – 20. The FOV and matrix size used for MRF and TMRF is 22.5cm2 and 225×225 respectively. Slice thickness and the number of slices were identical to the gold standard. The total scan time for the gold standard, MRF, and TMRF were approximately 22, 6, and 4 minutes respectively. The auto TI feature was selected for the T_1_ map sequence where there was modest variation for each slice as shown in a footnote. FSE – fast spin echo, FLAIR – fluid-attenuated inversion recovery, MRF – magnetic resonance fingerprinting, TMRF – tailored MRF, FOV – field of view, TR – repetition time, TE – echo time, TI – inversion time, FA – flip angle, RBW – receiver bandwidth, ETL – echo train length.

### *Qualitative imaging* studies

MRF and TMRF sequences leveraged an 89-shot spiral with a 608-point readout. The MRF and TMRF cases resulting in acquisition times were 5:57 (minutes: seconds) and 4:07, respectively. All sequences (GS, MRF, and TMRF) were acquired twice to compute the signal-to-noise ratio (SNR) using the difference image method [50]. Slice planning was maintained across all the sequences for each volunteer to allow spatial comparisons.

### TMRF reconstruction

The obtained raw data were pre-processed by compensating for a calibrated gradient delay (3.5μs), scaling k-space with the ratio of the field of view (FOV) to matrix size, removing spikes (threshold of twice the standard deviation of the FID), and weighting the k-space data with the pre-computed density compensation factor. We employed the non-uniform fast Fourier transform (NUFFT) to reconstruct the data [51], followed by complex coil combination and sliding window reconstruction to obtain 2D multi-slice images over time.

### MRF synthetic images

MRF provides T_1_ and T_2_ parametric maps simultaneously. Along with acquisition parameters such as TR, TE, and inversion time (TI), these maps were provided as inputs to the MR signal equation to synthetically generate multi-contrast contrast images such as T_1_ weighted, T_1_ FLAIR, and T_2_ weighted synthetic images.

### Deep learning (DL) based denoising

We denoised the TMRF images to account for the noise due to accelerated imaging [47]. The model was trained using the human connectome project (HCP) data. It comprised 8295 and 6622 T_1_ magnetization prepared - rapid gradient echo (MPRAGE) and T_2_ weighted images respectively. Forward modeling of noisy data involved obtaining noise patches from the noisy target data by cropping the four corners of the noisy images. Later, the noisy patches were accumulated and added to the HCP data with the image intensity level similar to the highest intensity level observed in the original TMRF data set. HCP datasets (clean and noisy) were used for training the native noise denoising network (NNDnet) employing a U-net. The rectified linear unit activation function was used, and the network was trained for 400 epochs on a computer with four Nvidia Tesla GPUs. The model was expected to denoise the TMRF images while simultaneously being aware of the signal and noise ratios in the test dataset. The T_1_ and T_2_ weighted images from the TMRF reconstruction were the test images. Gradient anisotropic diffusion (GAD) denoising available in the 3D Slicer was used as a comparative method to denoise the test images. We did not apply DL denoising to synthetically generated images from MRF as these images were computed from relaxometric maps rather than directly from k-space data. The noise, therefore, in these synthetic images arose from estimation errors in the quantitative maps rather than low SNR k-space data.

### Quantitative imaging studies - TMRF reconstruction and pattern matching

A DL approach based on DRONE [49] was used for TMRF quantitative tissue relaxometry map reconstruction. The architecture consisted of a four-layer, fully connected neural network. The input layer consisted of 749 nodes corresponding to the total number of time points. The output layer consisted of two nodes: T_1_ and T_2_. Hyperbolic tangent (tanh) was used as the activation function for the hidden layers and the sigmoid function for the output layer. Each of the hidden layers had 300 nodes. The network was trained using a learning rate of 0.001 and mean square error as the loss function. The network was trained for 500 epochs using an RMSprop optimizer. A total of 108808 samples were split into training, validation, and testing data with corresponding ratios of 0.71, 0.18, and 0.11, respectively.

The dictionary simulation was carried out using T_1_ in the range of 0 to 3000ms in increments of 2ms between 1 to 300ms and increments of 10ms between 300 to 3000ms. T_2_ was in the range of 0 to 1500ms in increments of 2ms between 1 to 300ms and increments of 10ms between 300 to 3000ms as in [49]. Entries with T_1_<T_2_ were excluded for physiological reasons. The absolute value of the dictionary entry was used as training data. The training required 95 minutes on an Nvidia 1060 (Nvidia Corp., Santa Clara, CA) GPU. This DRONE-like network was used to compute the quantitative T_1_ and T_2_ maps for both *in vivo* and *in vitro* data. The GS T_1_ curve fitting for *in vivo* brain data was performed offline in MATLAB, whereas T_2_ was obtained directly from the GE scanner’s software.

### MRF reconstruction and pattern matching

MRF reconstruction and pattern matching follow the same protocol as in [52] using the dictionary matching method.

### Image analysis

3D slicer [48] was used to segment GM and WM tissues in qualitative and quantitative data for all the slices and all four *in vivo* datasets. Skull stripping was performed manually. Tissue segmentation was performed semi-automatically using the threshold method in the 3D Slicer. The segmentation was performed on all the contrasts (except water and fat images) and two maps. The SNR was plotted and compared across all three methods. The mean contrast (the difference between WM and GM signal intensities) was calculated for all three methods and plotted using GraphPad Prism.

## RESULTS AND DISCUSSION

### Design and simulation

Figure 2 (a-c) depict the TE, FA, and TR train for MRF (dashed line) and TMRF (solid line). Figure 2 (d,e) shows the representative MRF and TMRF sliding window simulated signal evolutions for the four tissue types - WM (T_1_=860ms, T_2_=80ms for simulation), GM (T_1_=1320ms, T_2_=120ms), CSF (T_1_=4000ms, T_2_=1700ms), and fat (T_1_=380ms, T_2_=60ms). Figure 2 (e) depicts the TMRF signal evolutions that exhibit a slowly varying magnitude than MRF, ensuring better image reconstruction while using a sliding window. The subtler changes in TMRF signal evolutions increase challenges for quantitative imaging. However, DRONE and other DL-based methods are expected to be sensitive to these subtle changes. The inclusion of a 90^°^ pulse at the beginning of the second acquisition block introduced artifacts in sliding window reconstruction. This pulse was necessary to flip the magnetization to enable subsequent water and fat imaging. While the MRF sequence consists of 1000 time points, the TMRF sequence utilizes only 749 time points to acquire all five contrasts, resulting in a 25% reduction in scan time. TMRF takes a longer time for reconstruction (∼20 minutes for five qualitative images and two maps) than MRF (∼2 minutes for three synthetic contrasts and two maps). The longer reconstruction time is attributed to the sliding window reconstruction of all 749 time points to obtain natural contrast images. However, this process is performed only once on the first dataset to choose the time points corresponding to the five desired contrasts manually. These chosen time points were fixed for all subsequent datasets.

### Qualitative studies

Figure 3 shows the qualitative healthy brain images using the GS sequences, MRF, and TMRF in the first, second, and third rows, respectively. Each column represents different contrasts for the same subject. The window levels of the images were adjusted manually to depict good contrast.

**Figure 3:**
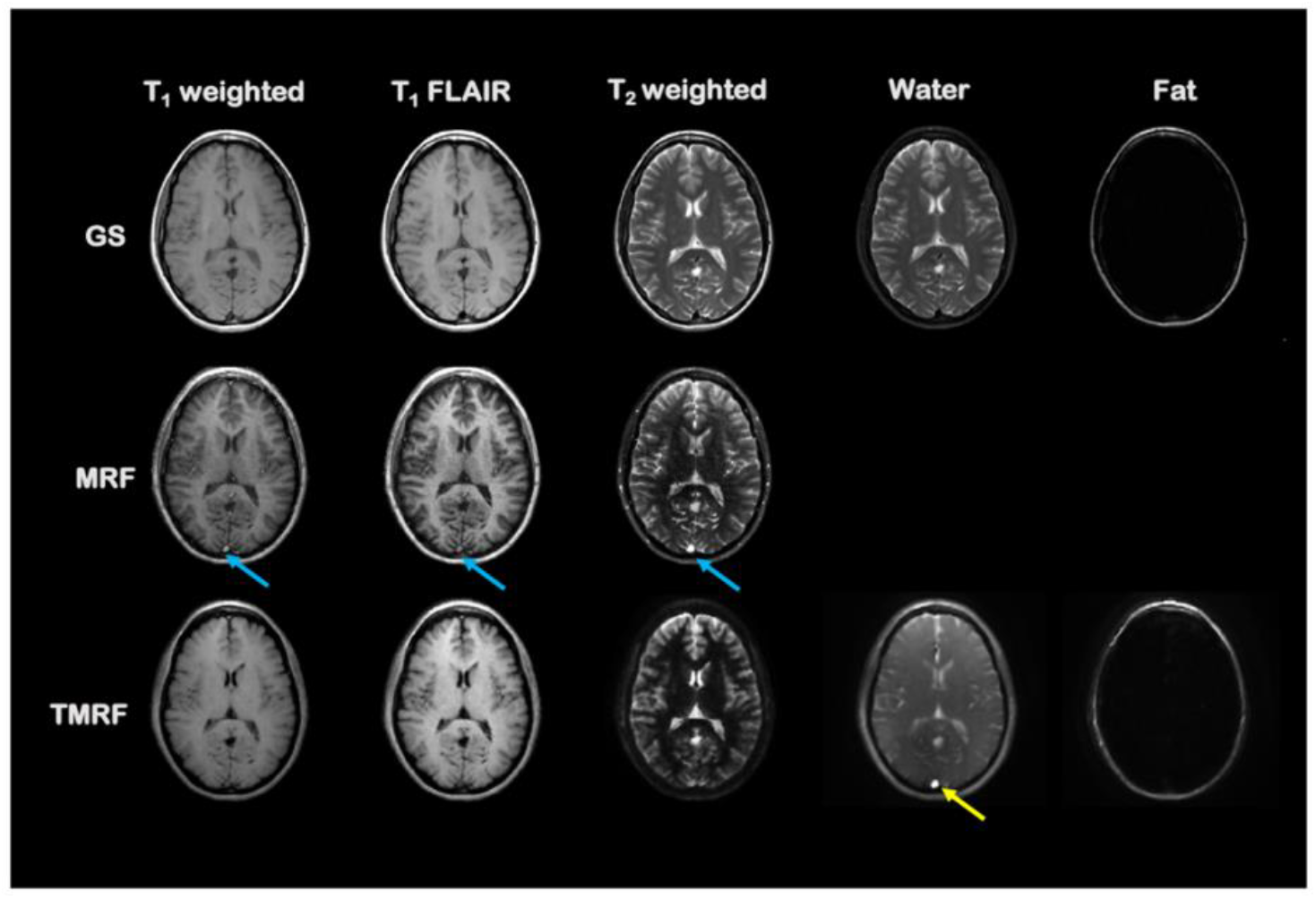
Qualitative studies: Qualitative healthy brain images obtained using GS method (first row), MRF (second row) and TMRF (third row). Window levels were manually adjusted to depict good contrast in the images. Each column represents different contrasts. GS, MRF, and TMRF were acquired twice to estimate the SNR by applying the “difference image” method. The same slice planning was maintained across all the sequences for each volunteer to facilitate spatial comparisons. For the first volunteer, five distinct contrast images were chosen from 749 reconstructed images. The time points 1, 95, and 150 were visually chosen for T_2_ weighted, T_1_ FLAIR, and T_1_ weighted respectively. For 2-point Dixon imaging, time points 575 and 675 were chosen. The same time point values were used for the other volunteers. Images obtained using MRF method were synthetically generated and show flow artifacts (blue arrows) in all the contrasts. Water images obtained from TMRF also show flow artifacts (yellow arrow); GS – gold standard, MRF - magnetic resonance fingerprinting, TMRF - tailored MRF, SNR - signal-to-noise ratio, FLAIR - fluid-attenuated inversion recovery.

All qualitative images obtained from TMRF are natural and show smooth transitions between GM and WM compared to MRF images. MRF images increased contrast between WM and GM as compared to GS and TMRF. However, these images are synthetically generated and are prone to partial volume artifacts and flow (indicated by blue arrows). TMRF water images suffer from flow artifacts, as indicated by the yellow arrow in Figure 3. The fat from the T_2_ weighted image of TMRF relatively shows a lesser signal when compared to GS and MRF as the time points selected for the T_2_ weighted images show a lesser fat signal, which is seen in simulation (Figure 2). Figure 4 shows the representative T_1_ weighted healthy brain images (first column) obtained from GS, MRF, and TMRF. The magnified images (second column) show that synthetically generated images exhibit partial volume effects resulting in patchy images (second row). We attribute this to the flow inherent in the Dixon window time points reflected in the sliding window reconstruction. The MRF (second row) and TMRF (third row) images are moderately rotated compared to the GS images depicting the subject motion between scans. TMRF can provide multiple images of the same contrast as for the first dataset. The time points for all the contrasts were selected manually. Hence, users can choose the best image for each contrast (Supplementary Figure 1). Once the time points were selected for the first dataset, the sliding window reconstruction was performed only on those selected time points for the remaining datasets (Figure 1). The water image computed from TMRF data appears to have flow and shading artifacts. We attribute this to imbalanced gradients and aim to compensate for flow during Dixon acquisitions in future implementations.

**Figure 4:**
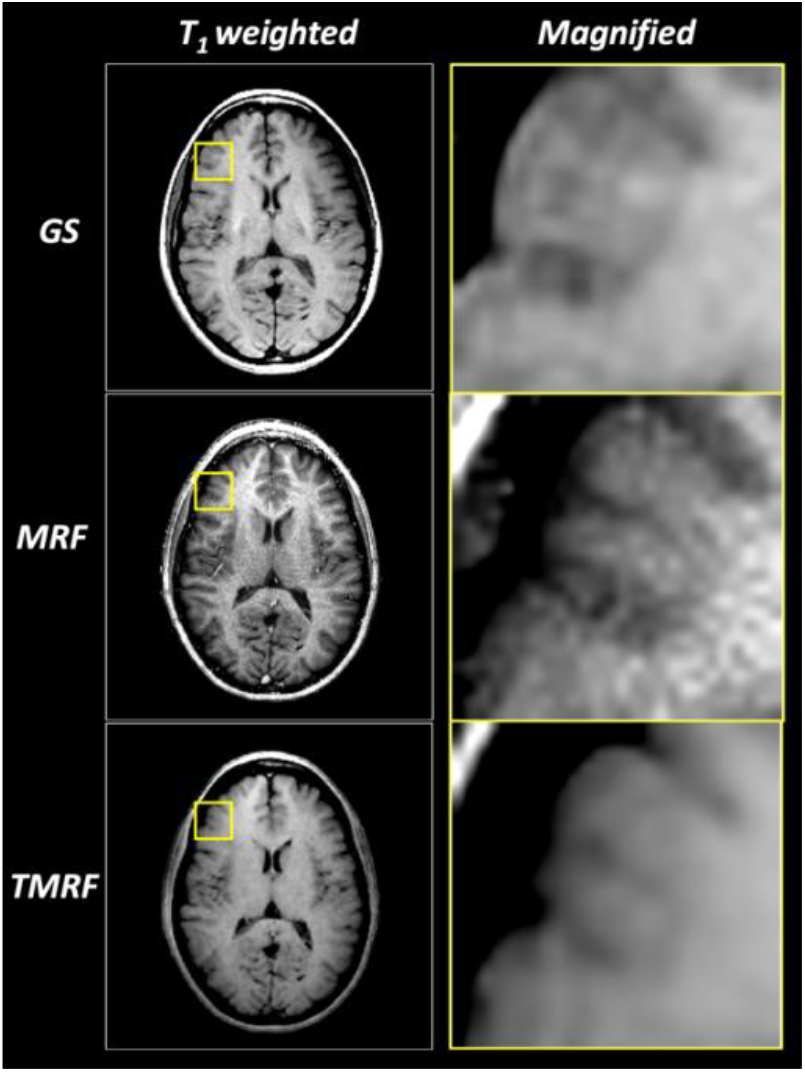
Magnified images: The representative T_1_ weighted and corresponding magnified images for all three methods (GS, MRF and TMRF) are shown in the first and second column respectively. The TMRF data shown here is post DL denoising. The yellow square box on the images shows the part which is magnified. These images are synthetically generated and are prone to flow artifacts, partial volume effects leading to patchy images. The GS, MRF and TMRF images show a difference in the rotation due to subject motion between these scans. Hence the part of the skull is seen in MRF and TMRF but not in GS; GS - gold standard; MRF - magnetic resonance fingerprinting; TMRF - tailored MRF; DL - deep learning.

### DL denoising

Supplementary Figure 2 shows the images of the representative denoised T_1_ weighted (a-d) and T_2_ weighted (e-h). The images denoised using NNDnet demonstrate a good balance between denoising and preserving edge features compared to the GAD filter. DL denoising was beneficial as the signal and noise levels were tailored to the noisy target image and preserved the native noise structure.

### Quantitative studies

Figure 5 presents the T_1_ and T_2_ quantitative maps of ISMRM/NIST phantom and healthy brains using the GS sequences, MRF, and TMRF in the first, second, and third rows, respectively. In Figure 5 (a), the quality of T_1_ maps of MRF and TMRF is similar to GS T_1_ maps. The T_2_ map of TMRF has increased artifacts compared to the T_2_ map of MRF. The artifacts observed in phantom and healthy brains may be attributed to B_1_ variations, as shown in Supplementary Figure 3. In Figure 5 (b), flow artifacts can be observed both in T_2_ maps of MRF and TMRF but not in the T_2_ map of GS due to the spiral trajectory without flow compensating gradients used in both MRF and TMRF. In Figure 5 (c), T_1_ estimates showed a strong linear correlation between TMRF and spin-echo (R^2^ = 0.9992), while the corresponding value for T_2_ was R^2^ = 0.9839. Figure 5 (d), T_1_ and T_2_ estimates also showed a strong linear correlation between MRF and spin-echo with R^2^ equal to 0.9965 and 0.9848, respectively.

**Figure 5:**
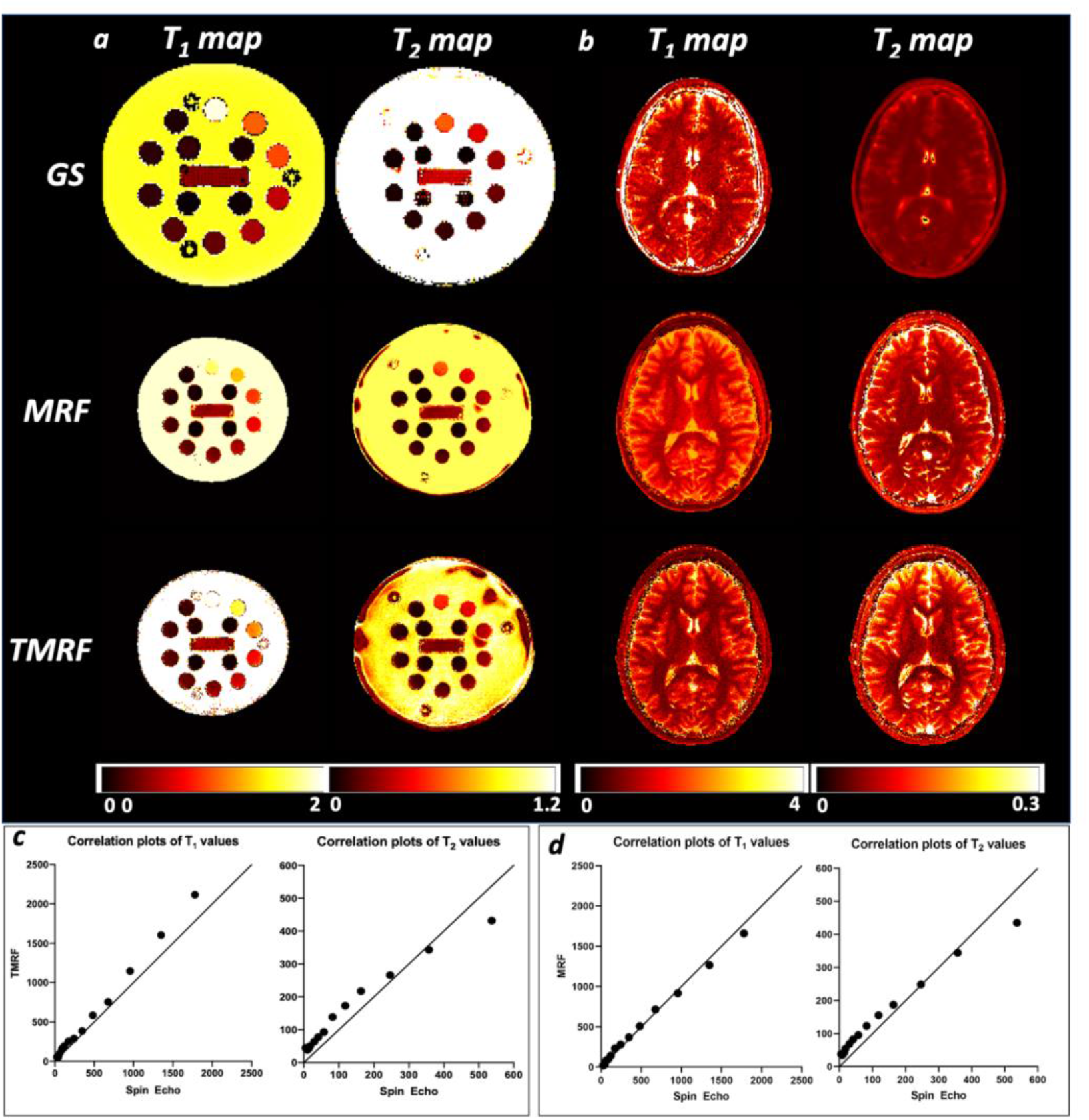
Quantitative studies: T_1_ and T_2_ maps of ISMRM/NIST phantom and brains of healthy human subjects as well as the correlation plots of T_1_ and T_2_ estimates of TMRF compared to the spin-echo methods. a) T_1_ and T_2_ maps of ISMRM/NIST phantom. In the first row, T_1_ values were estimated from the T_1_ array using the IR-SE method, and T_2_ values were estimated from the T_2_ array using the SE method. In the second row, T_1_ values and T_2_ values were estimated using the MRF method. In the third row, T_1_ values and T_2_ values were estimated using the TMRF method. b) T_1_ and T_2_ maps of healthy human subjects’ brains using GS, MRF and TMRF methods. c) T_1_ and T_2_ estimates of TMRF compared to the spin-echo methods. d) T_1_ and T_2_ estimates of MRF compared to the spin-echo methods. The solid line depicts a straight line with a slope equal to 1; IR-SE – inversion recovery - spin echo, GS – gold standard, MRF - magnetic resonance fingerprinting, TMRF - tailored MRF.

### Image analysis

Figure 6 (a) shows the plot of SNR (first row) and signal intensity (second row) of WM and GM of qualitative data for all the three methods (GS, MRF, and TMRF) and three contrasts (T_1_ weighted, T_1_ FLAIR, and T_2_ weighted) for four subjects. Figure 6 (b) shows the plot of T_1_ and T_2_ values of WM and GM for T_1_ and T_2_ maps and all three methods. Supplementary Figure 4 shows a healthy brain’s representative GS T_1_ weighted image and its corresponding GM and WM segmented mask using the 3D slicer software. The GS images show better SNR in both WM and GM for all three contrasts and all four subjects than MRF and TMRF (Figure 6 (a)). TMRF shows better SNR than MRF for T_1_ weighted images and T_1_ FLAIR, whereas TMRF and MRF are similar for T_2_ weighted images for all four datasets. The T_2_ weighted image obtained from TMRF is noisy as the signal intensity during that time is lower, as seen in the magnetization evolution (Figure 2). The second row of Figure 6 (a) depicts the average intensity values of WM and GM for TMRF and are relatively higher compared to GS and MRF for T_1_ weighted and T_1_ FLAIR images. For MRF, the average intensity values for WM and GM are lower compared to GS and TMRF. This may be attributed to the differences in imaging parameters (TR and TE) and the reconstruction method. The T_1_ and T_2_ values of WM and GM of MRF data for all the four subjects are relatively overestimated, compared to GS and TMRF (as seen in Figure 6 (b)). The GS and TMRF values are within a similar range. The segmented GM and WM relaxation time values are the average values of the entire segmented area. This averaging results in loss of spatial localization and broader ranges of the values. TMRF provides five contrasts and two maps in 4:07 (minutes: seconds), whereas GS takes 22:10 (min: sec) for the same number of images, and MRF takes 5:57 (min: sec) for three contrasts and two maps. Scan efficiency for TMRF is higher (1.72 min^-1^) than GS and MRF, which correspond to 0.32 min^-1^ and 0.90 min^-1^, respectively. MRF can theoretically produce an infinite number of contrasts synthetically. However, these images are corrupted by flow and partial volume artifacts (see Figures 3 & 4). This is because it is numerically challenging to include a complete simulation of physical effects such as flow, diffusion, multi-component voxels, etc.

**Figure 6:**
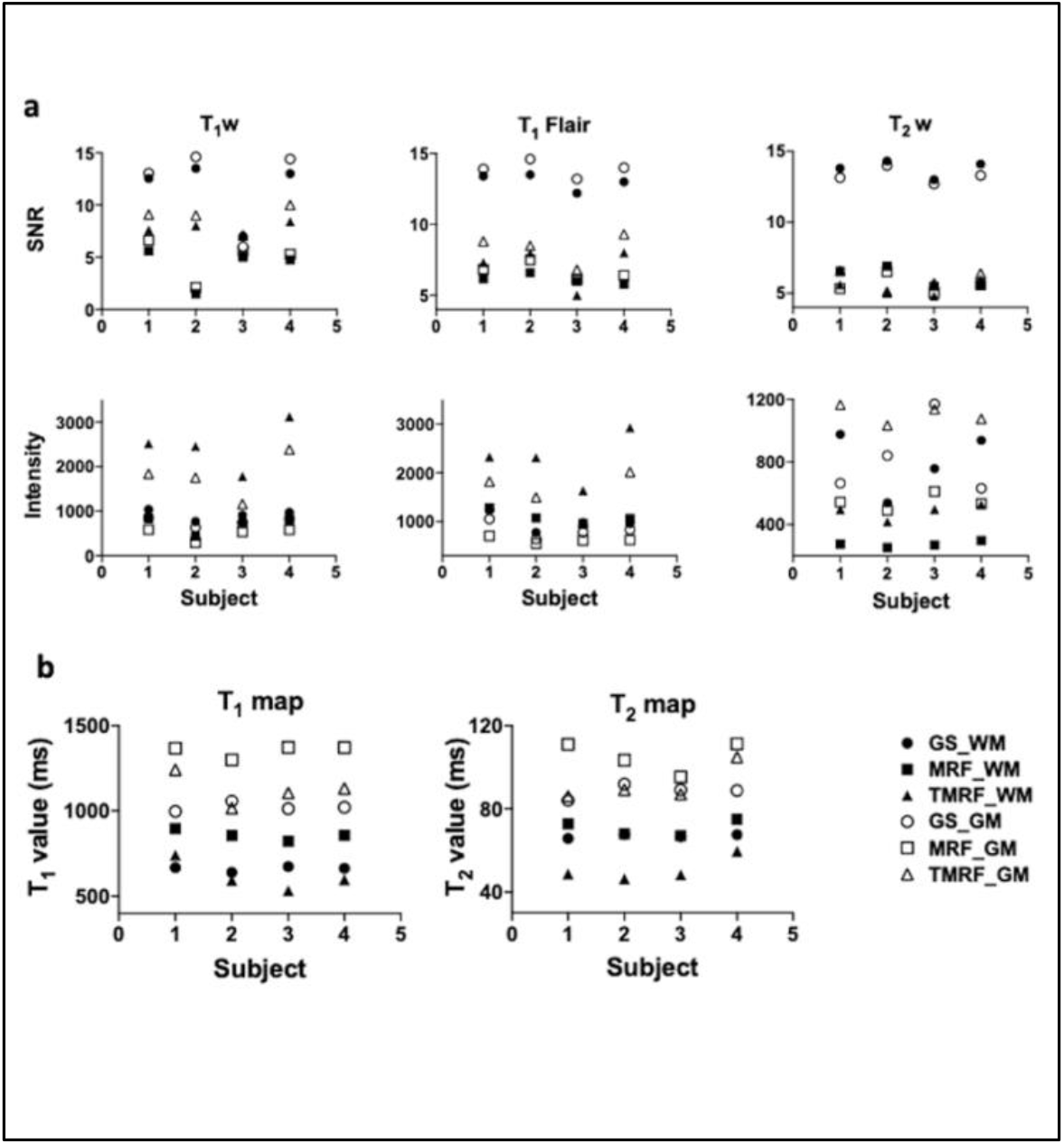
Image analysis: (a) The first row shows a plot of SNR, the second row shows the signal intensity of WM and GM of qualitative data of four subjects for GS, MRF, and TMRF, for three contrasts (T_1_ weighted, T_1_ FLAIR, and T_2_ weighted). For all the three contrasts across four subjects, GS images show better SNR in both WM and GM, compared to MRF and TMRF. For T_1_ weighted image and T_1_ FLAIR, TMRF presents a better SNR. However, for T_2_ weighted images, TMRF and MRF are similar for all four datasets. (b) shows the plot of T_1_ and T_2_ values of WM and GM for three different contrasts and all three methods. For T_1_ weighted and T_1_ FLAIR images, the average intensity values of WM and GM for TMRF (second row) are comparatively higher compared to GS and MRF.; SNR - signal to noise ratio, WM - white matter, GM - grey matter, GS - gold standard, MRF - magnetic resonance fingerprinting, TMRF - tailored MRF, FLAIR - fluid-attenuated inversion recovery.

We have demonstrated rapid acquisition of simultaneous, natural (non-synthetic), five contrasts, and two quantitative maps in this work. In particular, TMRF requires approximately four minutes compared to the GS sequences requiring twenty-five minutes. This will accelerate imaging of pathologies such as brain tumors and multiple sclerosis (refer to Introduction section), especially in pediatric populations. The low SNR contrast (T_2_ weighted) in TMRF leveraged DL denoising. The need for denoising can be gleaned from TMRF’s design and simulation outcomes. TMRF’s design is flexible, and future implementations can incorporate other contrasts such as DWI [53] and T_2_ FLAIR [54]. The choice of FA, TR, and TE in this study was based on knowledge of the tissue matters and contrasts for short TRs. This design can be extended to image other tissue types and organs.

## Data Availability

All data produced in the present study are available upon reasonable request to the authors

## ACKNOWLEDGEMENT

We would like to acknowledge Rolf F. Schulte for providing multi nuclear spectroscopy research pack (MNSRP); Guido Buonincontri and Pedro A Gomez for MRF package on MNSRP.

