## Supplementary materials for "Tailored Magnetic Resonance Fingerprinting"

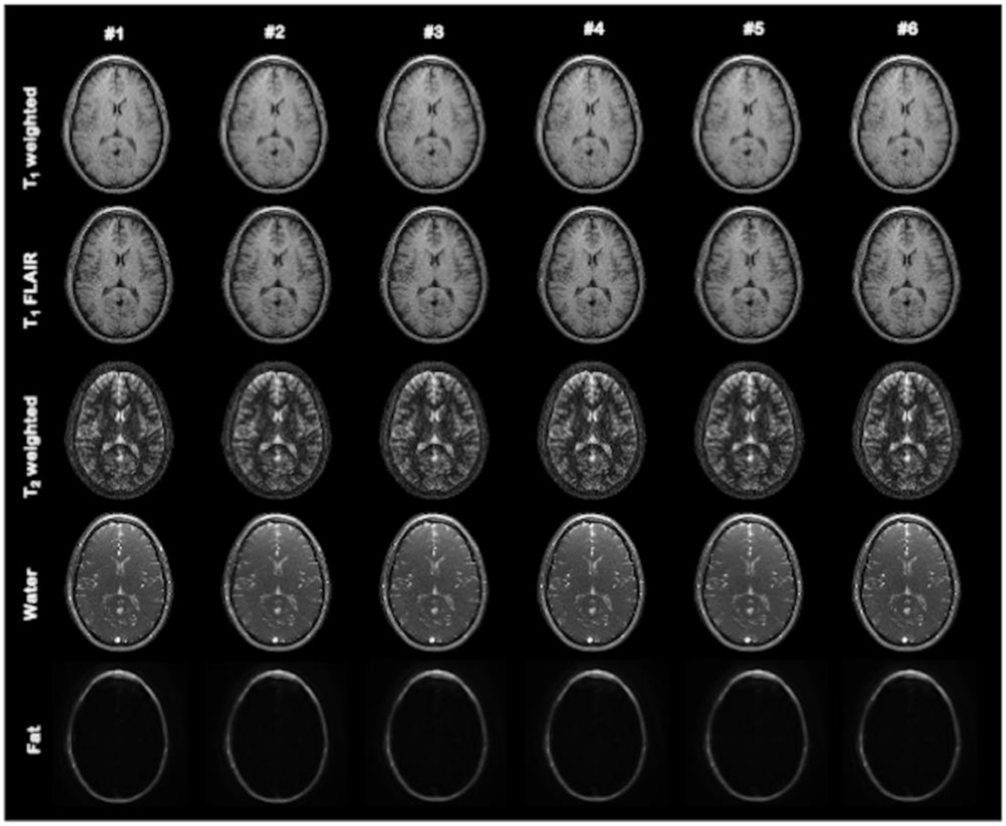


**Supplementary Figure 1: One time procedure to pick time-points:** Shows the TMRF reconstructed images with six images (column) for all five contrasts (rows). The time points for each contrast were selected based on visual inspection. Hence "length of contrast window" is given as an input. Here, the window size is 6 where (timepoint - 3) and (timepoint +2) images were reconstructed. Users can choose the best image (timepoint) from each of the rows (contrast). This step was carried out only once for the first dataset and the sliding window reconstruction was performed on the selected time points for the rest of the datasets. All images shown here are before DL denoising; TMRF - tailored MRF, DL - deep learning, FLAIR - fluid-attenuated inversion recovery.

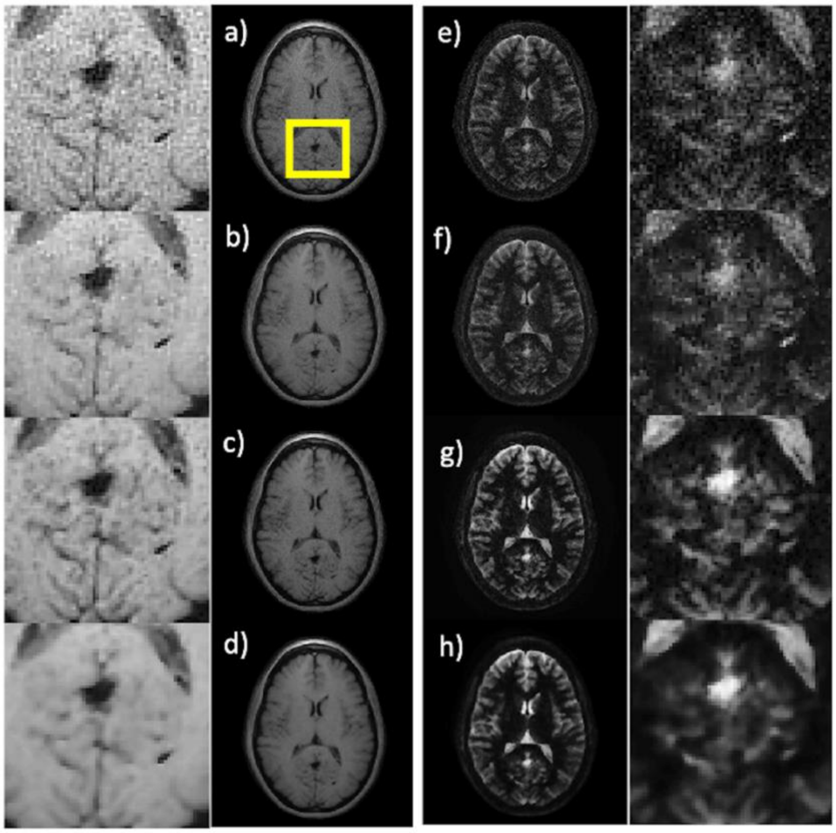


**Supplementary Figure 2: DL denoising:** Denoising $T_1$ weighted images (a-d) - a) a $T_1$-weighted image from TMRF; b) denoised using the GAD filter; c) corresponding NNDnet result and d) denoising combining NNDnet and GAD. The first column shows the magnified images for the yellow square shown in a). Corresponding denoised $T_2$-weighted images and their magnified portions are shown in (e-h) and the fourth column. NNDnet images show a good balance between denoising and preserving edge features; TMRF - tailored MRF, GAD - gradient anisotropic diffusion, NNDnet - native noise denoising network.

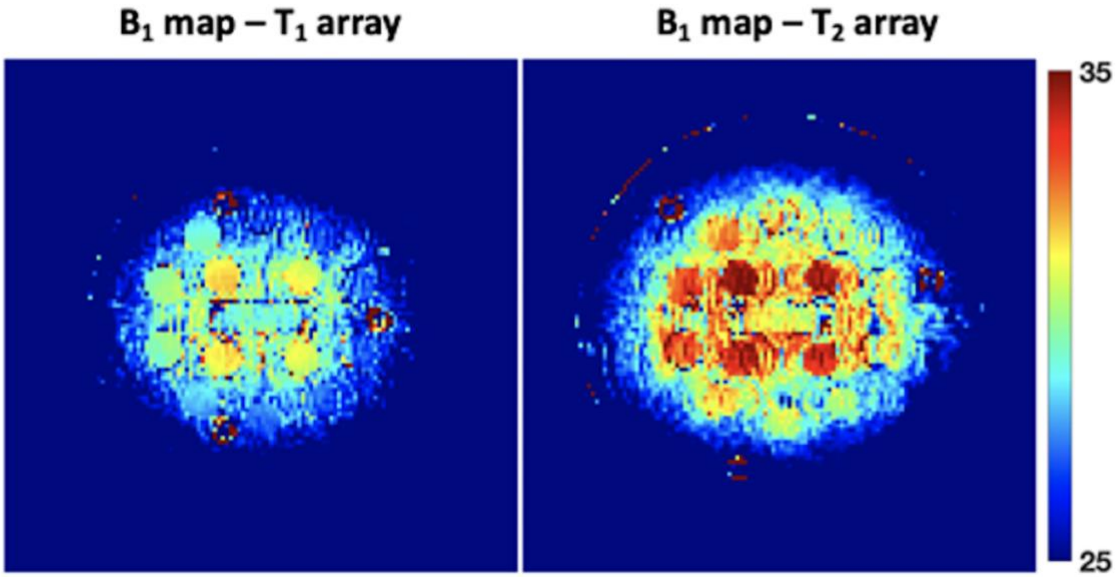


**Supplementary Figure 3: $B_1$ map:** $B_1$ map of $T_1$ and $T_2$ array of ISMRM/NIST phantom. The map was acquired using the Bloch-Siegert method with a target flip angle of 30 degrees. The four spheres with small $T_2$ values experience higher flip angles compared to other spheres representing the $B_1$ inhomogeneity in the data. The GS relaxometry measurements of the ISMRM/NIST phantom are shown in supplement table 1; ISMRM/NIST - International Society for Magnetic Resonance in Medicine/National Institute of Standards and Technology, GS – gold standard.

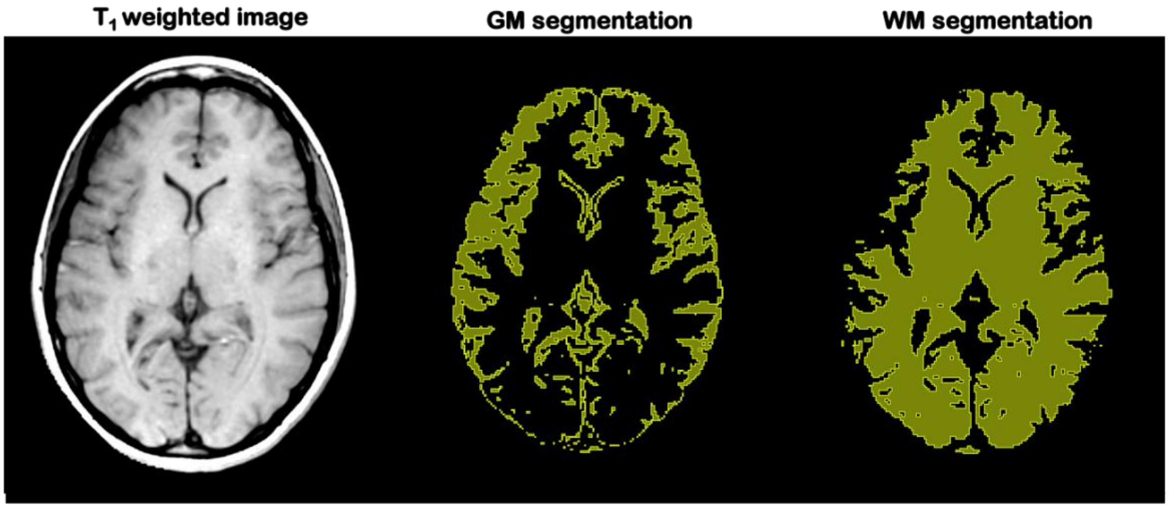


**Supplementary Figure 4: Segmentation:** Representative $T_1$ weighted healthy human brain image along with GM and WM segmentation. The segmentation was performed with the help of Slicer 3D software using threshold-based method. Skull stripping was performed before the segmentation; GM – grey matter; WM – white matter.

| | | 1 | 2 | 3 | 4 | 5 | 6 | 7 | 8 | 9 | 10 | 11 | 12 | 13 | 14 |
|---|---|---|---|---|---|---|---|---|---|---|---|---|---|---|---|
| $T_1$ (ms) | Mean | 1780 | 1351 | 957 | 678 | 483 | 346 | 242 | 173 | 120 | 90 | 60 | 44 | 32 | 24 |
| | SD | 44 | 17 | 6 | 82 | 47 | 4 | 3 | 17 | 15 | 11 | 8 | 1 | 1 | 2 |
| $T_2$ (ms) | Mean | 537 | 357 | 246 | 163 | 118 | 82 | 57 | 41 | 30 | 18 | 14 | 10 | 8 | 5 |
| | SD | 10 | 5 | 5 | 2 | 1 | 1 | 1 | 1 | 3 | 1 | 1 | 1 | 1 | 1 |

**Supplementary Table 1: $T_1$ and $T_2$ of NIST phantom:** Gold standard relaxometry measurements of the ISMRM/NIST phantom in a wide bore scanner. The mean and SD of $T_1$ estimates of the 14 spheres from the $T_1$ array using the IR-SE method and $T_2$ estimates of 14 spheres from the $T_2$ array using the spin-echo method; ISMRM/NIST - International Society for Magnetic Resonance in Medicine/National Institute of Standards and Technology, SD - standard deviation, IR-SE – inversion recovery - spin echo.
